# Incidence and progression of diabetic retinopathy and blindness in Indonesian adults with type 2 diabetes

**DOI:** 10.1101/2025.03.18.25324213

**Authors:** Muhammad B Sasongko, Gandhi A Febryanto, Supanji, Firman S Wardhana, Yeni D Lestari, Anggraeni Puspita, Felicia Widyaputri

## Abstract

**Objectives:** To report the incidence and progression rate of diabetic retinopathy (DR) and blindness in Indonesian adults with type 2 diabetes.

**Methods:** This was a prospective cohort study of 899 adults aged >30 years with confirmed type 2 diabetes. All participants underwent standardized clinical and eye examinations. Two-field retinal photographs were taken. DR was graded by trained grader masked to participants’ clinical details. We categorized DR as follows: mild, moderate, severe non-proliferative (NPDR), and proliferative DR (PDR). Additional category of vision-threatening DR (VTDR) included severe NPDR or more, or moderate NPDR with clinically significant macular edema. Blindness was defined as visual acuity ≤3/60. At least 1-step progression was categorized as DR progressing. Cox-proportional hazard model was used.

**Results:** The incidence and progression of DR were 34.6 and 35.1, and incidence of VTDR and blindness were 24.5 and 8.33/1000 person-years, respectively. Longer diabetes duration was associated with increasing the risk of developing DR (Hazard ratio 1.37 [95% confidence interval 1.10-1.72]) and VTDR (2.00 [1.60-2.50]) and progressing DR (1.50 [1.23-1.84]) in 5 years. Obesity was associated with increased risk of developing DR (1.75 [1.14-2.68]) and the presence of gangrene (2.54 [1.49-4.35]) and neuropathy (1.50 [1.07-2.10]) at baseline increased the risk of progressing DR in 5 years. Living in rural area was associated with increased risk of blindness (2.50 [1.03-5.88]).

**Conclusions:** We reported the incidence and progression rate of DR, VTDR, and blindness, and documented that longer diabetes duration increased the risk of DR and VTDR in 5 years.

## Introduction

Diabetic retinopathy (DR) is the most frequent microvascular complication in persons with diabetes and the leading cause of visual impairment and blindness among working age population [1,2]. Diabetes has become a significant public health problem in developing countries particularly in Asia [3–5]. Worldwide, the number of people with diabetes is constantly growing, and in Indonesia, it is estimated to reach over 20 million in 2030, ranked as the fourth largest diabetes population in the world [3–5].

With increasing number of diabetes, so is presumed the number of DR. Data from our previous study has documented that the prevalence of DR and DR-related blindness in Indonesia is higher than other countries [6]. However, we have not yet reported the incidence and progression of DR in our population. To date, published studies that report incidence and progression of DR from developing countries are very limited [7]. Such data are fundamental for priority setting to develop national plan and public health strategy to reduce diabetes-related blindness. In this study, we aimed to report the incidence and progression rate of DR and VTDR, and the incidence of DR-related blindness in Indonesia.

## Methods

### Study population

This was a prospective cohort study, a 5-year follow-up of the Jogjakarta Eye Diabetic Study in the Community (JOGED.COM Study). JOGED.COM is the only community-based study of diabetes in Indonesia that recruited individuals aged ≥ 30 years with type 2 diabetes in 2014 – 2016 [6,8]. At baseline, we used a two-stage cluster random sampling method and recruited 1184 type 2 diabetes patients randomly from 23 community-health-centres (CHCs) in Jogjakarta [6,8]. This study began on November 1, 2019, and concluded on July 31, 2022. In this study, we randomly selected sub-sample of 900 individuals from the total cohort (N=1184) due to COVID-19 restriction. We excluded clusters with least number of baseline participants or with high cases of COVID-19. This study adhered to the principles of the Declaration of Helsinki. The ethical clearance, information sheet and consent form were approved by the Medical and Health Ethics Research Committee, Faculty of Medicine, Public Health, and Nursing, Unversitas Gadjah Mada (Ethics approval no. KE/FK/0431/EC/2019). Written informed consent was obtained from all participants.

### Clinical examinations, retinal photography and diabetic retinopathy assessment

Detailed methodology was identical to our baseline study [6,8,9]. Briefly, we performed standardized clinical and eye examinations including visual acuity (VA), external eye condition and thorough interview to assess self-reported risk factors. Two-field (optic disc and macula-centred) fundus photography was obtained using Kowa NonMyd-7 (Kowa, Japan) attached with Nikon D7000 camera. DR was graded from digital retinal photographs by trained grader masked to participants’ clinical details, side by side with baseline images to assessed the difference.

Eye-specific DR severity level was graded according to the modified Airlie House Classification system, and categorized as follows: ‘No DR’ was equal to Early Treatment for Diabetic Retinopathy Study (ETDRS) level 10 and 12, mild non-proliferative DR (NPDR) as ETDRS level 14 to 20, moderate NPDR as level 31 and 41, and severe NPDR and proliferative DR (PDR) as 51 to 80 [10]. Any DR was defined as levels 14 to 80 [11–13]. Clinically significant macular edema (CSME) was defined by the presence of retinal hard exudates within the central fovea area. Vision Threatening DR (VTDR) was defined to include severe NPDR, PDR and/ or CSME [11,13].

We defined several outcomes as follows: 1) incidence of DR – presence of any DR at the 5-year follow-up in individuals without DR at baseline; 2) incidence of VTDR – presence of VTDR at 5-year follow-up in individuals without VTDR at baseline; 3) DR progression – a 1 or more step increase in DR severity at 5-year follow-up in eyes with any DR at baseline; and 4) incidence of blindness was defined if an individual had VA ≥ 4/60 at baseline but reduced to ≤ 3/60 at the follow-up [14].

We performed statistical analyses using Stata BE 17 for Mac (Stata Corp., TX). Incidence and progression of DR, VTDR, and blindness were analysed using Cox-proportional Hazard regression model, adjusted for potential covariates. We presented the incidence and progression rates as the number of events per 1000 person-years.

## Results

Of 899 invited participants, 695 completed the examinations (77.9%) with a median follow-up time of 5.04 years (range 4.1-7.2). At baseline, there were 377 subjects without DR, 318 subjects with DR, 507 subjects without VTDR, and 520 subjects without blindness (**Fig 1**). Incidence and progression of DR, VTDR and blindness and their participants’ baseline characteristics is shown in Table 1. The overall incidence and progression rate of DR, incidence of VTDR and blindness were 34.6, 35.1, 24.5, and 8.3/1000 person-year, respectively. Participants who developed DR and VTDR at 5 years were diagnosed with diabetes at a younger age than those who were not (P=0.008 for incident DR and P<0.001 for incident VTDR). Those who developed DR and VTDR and whose DR was progressed had longer diabetes duration at baseline than those who did not (P<0.001 for incident DR and VTDR and progression of DR). Participants who became blind after 5 years had higher systolic blood pressure (SBP) (P=0.037) and more likely to be resident of rural areas (P=0.024) compared to those who did not. On the contrary, in the subjects with no blindness at baseline, the SBP (P=0.037) and location of residence (P=0.024) were significantly different between those who developed blindness at follow-up and those who did not.

**Table 1.** Baseline characteristics of participants by incident DR, progressed DR, incident VTDR and blindness.

|  | No DR at Baseline<br>(N= 377) |  |  | Any DR at Baseline<br>(N=318) |  |  | No VTDR at Baseline (N=507) |  |  | No Blindness at Baseline<br>(N= 520) |  |  |
| --- | --- | --- | --- | --- | --- | --- | --- | --- | --- | --- | --- | --- |
|  | No DR at 5-year | Any DR at<br>5-year | P-value | DR stable at<br>5-year | DR<br>worsened at<br>5-year | P-value | No VTDR at<br>5-year | VTDR at 5-year | P-value | No blindness<br>at 5-year | Blindness<br>at 5-year | P-value |
| Total subject | 235 | 142 | - | 175 | 143 | - | 406 | 101 | - | 486 | 34 | - |
| Incidence/<br>Progression rate | 34.6/1000 person-year (29.3-40.8) |  |  | 35.1/1000 person-year (29.8-41.3) |  |  | 24.1/1000 person-year (20.2-29.8) |  |  | 8.3/1000 person-year (5.9-11.7) |  |  |
| Gender* |  |  |  |  |  |  |  |  |  |  |  |  |
| Male | 61 (26) | 39 (27.5) | 0.75 | 55 (31.4) | 40 (28) | 0.50 | 106 (26.1) | 31 (30.7) | 0.35 | 131 (27) | 12 (35.3) | 0.29 |
| Female | 174 (74) | 103 (72.5) |  | 120 (68.6) | 103 (72) |  | 300 (73.9) | 70 (69.3) |  | 355 (73.0) | 22 (64.7) |  |
| Age at baseline | 56.4 ± 10.9 | 57.5 ± 10.5 | 0.34 | 57.6 ± 9.3 | 58.9 ± 8.5 | 0.19 | 57.0 ± 10.4 | 57.7 ± 10.6 | 0.53 | 56.9 ± 9.8 | 58.9 ± 8.9 | 0.25 |
| Age at diagnosis | 52.6 ± 10.9 | 49.5 ± 10.8 | **0.008** | 51.4 ± 9.8 | 50.6 ± 9.0 | 0.46 | 52.4 ± 10.1 | 47.7 ± 11.7 | **<0.001** | 51.2 ± 9.9 | 51.7 ± 10.2 | 0.81 |
| DM Duration** | 3 (1-5) | 8 (2-12) | **<0.001** | 4(2-8) | 7(3-12) | **<0.001** | 3 (1-7) | 10 (4-14) | **<0.001** | 4(2-8) | 7(2-10) | 0.06 |
| Fasting glucose<br>(mg/dL) | 202 (94.2) | 217 (99.2) | 0.17 | 210 (93.6) | 228 (115) | 0.13 | 202 (98.9) | 216 (98.2) | 0.28 | 211 (92.5) | 213 (111) | 0.92 |
| Systolic (mmHg) | 130.9 ± 20.6 | 130.8 ± 18.8 | 0.95 | 133.5 ± 20.7 | 135.0 ± 19.4 | 0.51 | 131.1 ± 19.9 | 131.5 ± 17.3 | 0.84 | 131.5 ± 19.4 | 138.8 ± 21.8 | **0.037** |
| Diastolic (mmHg) | 80.3 ± 12.2 | 81.1 ± 8.8 | 0.92 | 81.8 ± 8.4 | 82.6 ± 8.5 | 0.36 | 80.6 ± 11.3 | 82.2 ± 8.9 | 0.20 | 80.9 ± 10.9 | 81.8 ± 10.4 | 0.63 |
| BMI* |  |  |  |  |  |  |  |  |  |  |  |  |
| Under/<br>Normo-weight | 81 (34.4) | 42 (29.5) | 0.50 | 67 (38.3) | 65 (45.4) | 0.34 | 133 (32.8) | 40 (39.6) | 0.22 | 165 (34.0) | 17 (50) | 0.17 |
| Overweight | 100 (42.5) | 61 (43) |  | 68 (38.8) | 53 (37.1) |  | 175 (43.1) | 44 (43.6) |  | 208 (42.8) | 11 (32.3) |  |
| Obese | 54 (23) | 39 (27.5) |  | 40 (22.8) | 25 (17.5) |  | 98 (23.1) | 17 (16.8) |  | 113 (23.2) | 6 (17.6) |  |
| Education Level* |  |  |  |  |  |  |  |  |  |  |  |  |
| No School | 23 (9.8) | 13 (9.2) | 0.34 | 19 (10.8) | 19 (13.3) | 0.82 | 40 (9.8) | 12 (11.9) | 0.14 | 47 (9.7) | 5 (14.7) | 0.55 |
| Elementary | 86 (36.6) | 39 (27.5) |  | 62 (35.4) | 44 (30.8) |  | 140 (34.5) | 22 (21.8) |  | 157 (32.3) | 13 (38.2) |  |
| Junior HS | 100 (42.6) | 71 (50.0) |  | 77 (44.0) | 63 (44.0) |  | 182 (44.8) | 52 (51.5) |  | 224 (46.1) | 14 (41.2) |  |
| Senior HS | 25 (10.6) | 16 (11.3) |  | 17 (9.7) | 15 (10.5) |  | 41 (10.1) | 12 (11.9) |  | 54 (11.1) | 2 (5.9) |  |
| Smoking Status* |  |  |  |  |  |  |  |  |  |  |  |  |
| Not a Smoker | 169 (71.9) | 108 (76.1) | 0.57 | 127 (72.6) | 104 (72.7) | 0.94 | 295 (72.7) | 74 (73.3) | 0.95 | 356 (73.2) | 29 (85.3) | 0.34 |
| Active Smoker | 23 (9.8) | 12 (8.4) |  | 19 (10.9) | 14 (9.8) |  | 40 (9.8) | 11 (10.9) |  | 47 (9.7) | 2 (5.8) |  |
| Ex-smoker | 42 (17.9) | 20 (15.5) |  | 28 (16.0) | 24 (16.8) |  | 67 (16.5) | 16 (15.8) |  | 78 (16.0) | 3 (8.8) |  |
| Medication* |  |  |  |  |  |  |  |  |  |  |  |  |
| No Medication | 10 (4.2) | 6 (4.2) | 0.61 | 4 (2.3) | 4 (3.0) | 0.19 | 13 (3.2) | 4 (4.0) | 0.07 | 11 (2.26) | 1 (2.9) | 0.83 |
| Oral Medication | 194 (82.5) | 113 (79.6) |  | 144 (82.3) | 105 (73.4) |  | 340 (83.7) | 76 (75.2) |  | 403 (82.9) | 27 (79.4) |  |
| Oral + Insulin | 28 (11.9) | 22 (15.4) |  | 27 (15.4) | 33 (23.1) |  | 49 (12.1) | 21 (20.8) |  | 69 (14.2) | 6 (17.6) |  |
| Stroke and CVD* |  |  |  |  |  |  |  |  |  |  |  |  |
| Yes | 10 (4.2) | 7 (4.9) | 0.76 | 9 (5.1) | 4 (3.0) | 0.30 | 17 (4.1) | 5 (4.9) | 0.75 | 21 (4.3) | 1 (2.9) | 0.69 |
| No | 223 (94.9) | 134 (94.4) |  | 166 (94.8) | 138 (96.5) |  | 385 (94.2) | 96 (95.1) |  | 462 (95.1) | 33 (97.0) |  |
| Gangrene* |  |  |  |  |  |  |  |  |  |  |  |  |
| Yes | 6 (2.5) | 7 (4.9) | 0.22 | 16 (9.1) | 16 (11.2) | 0.53 | 11 (2.7) | 9 (8.9) | **0.005** | 25 (5.1) | 4 (11.8) | 0.53 |
| No | 227 (96.6) | 134 (94.4) |  | 159 (90.8) | 126 (88.1) |  | 391 (96.3) | 92 (91.1) |  | 458 (94.2) | 30 (88.2) |  |
| Neuropathy* |  |  |  |  |  |  |  |  |  |  |  |  |
| Yes | 105 (44.7) | 70 (49.3) | 0.39 | 102 (58.3) | 89 (62.2) | 0.43 | 191 (47.0) | 54 (53.5) | 0.29 | 229 (47.1) | 16 (47.0) | 0.97 |
| No | 128 (54.5) | 71 (50.0) |  | 73 (41.7) | 53 (37.1) |  | 211 (52.0) | 47 (46.5) |  | 254 (52.3) | 18 (52.9) |  |
| Residence* |  |  |  |  |  |  |  |  |  |  |  |  |
| Rural | 159 (67.6) | 83 (58.4) | 0.07 | 125 (71.4) | 95 (66.4) | 0.34 | 268 (66.0) | 58 (57.4) | 0.11 | 307 (63.2) | 28 (82.3) | **0.024** |
| Urban | 76 (32.3) | 59 (41.5) |  | 50 (28.6) | 48 (33.6) |  | 138 (34.0) | 43 (42.6) |  | 179 (36.9) | 6 (17.6) |  |

All data are in mean (standard deviation) except indicated.
\*Median (inter-quartile range)
Abbreviations: BMI, body mass index; CVD, cardiovascular diseases; DM, diabetes mellitus; DR, diabetic retinopathy; HS, high school; VTDR, vision threatening diabetic retinopathy

**Figure 1.**
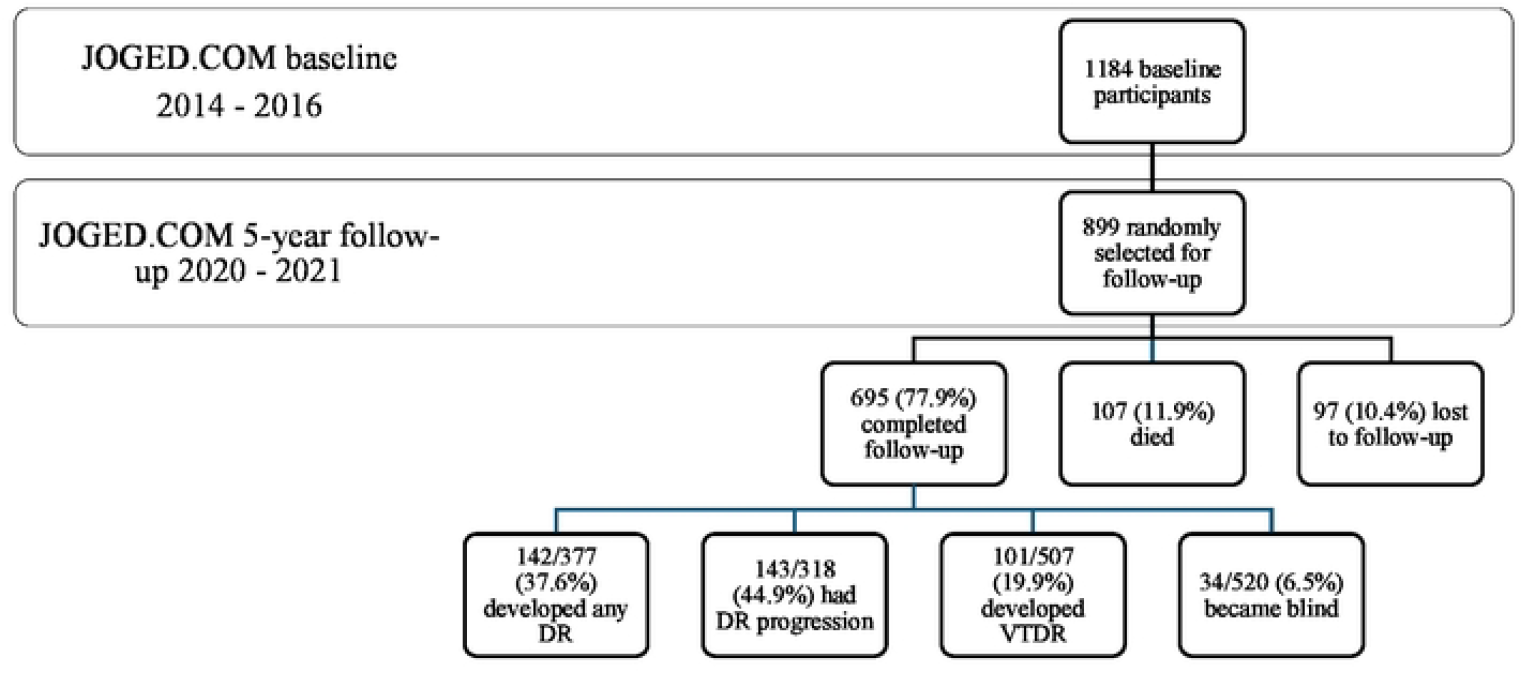
Flow of patient selection in JOGED.COM 5-year follow-up.

Table 2 shows the risk factors for DR and VTDR incidence and DR progression. Every 10-year increase in diabetes duration was significantly associated with increased risk of developing DR (Hazard ratio [HR] 1.37; 95% confidence interval [CI] 1.10-1.72; P=0.03) and VTDR (HR 2.00; 95% CI 1.60-2.50; P<0.01) and progressing DR (HR 1.50; 95% CI 1.23-1.84; P<0.01) in 5 years. Obesity was associated with increased risk of developing DR (HR 1.75; 95% CI 1.14-2.68; P=0.01 for obesity vs. normo-weight) and the presence of gangrene (HR 2.54; 95% CI 1.49-4.35; P<0.01) and neuropathy (HR 1.50; 95% CI 1.07-2.10; P=0.04) at baseline increased the risk of progressing DR in 5 years (**S1 Table**). In addition, living in rural area was associated with increased risk of blindness (HR 2.50; 95% CI 1.03-5.88; P=0.037) (**S2 Table**).

**Table 2.** Diabetic retinopathy incidence, VTDR incidence, and DR progression by gender, age at diagnosis, and diabetes duration.

| Risk Factors | DR Incidence |  |  |  |  | VTDR Incidence |  |  |  |  | DR Progression |  |  |  |  |
| --- | --- | --- | --- | --- | --- | --- | --- | --- | --- | --- | --- | --- | --- | --- | --- |
|  | At Risk | Incidence rate (95% CI) | HR (95% CI) | P<sup>1</sup> | P<sup>2</sup> | At Risk | Incidence rate (95% CI) | HR (95% CI) | P<sup>1</sup> | P<sup>2</sup> | At Risk | Progression rate (95% CI) | HR (95% CI) | P<sup>1</sup> | P<sup>2</sup> |
| Total subject | 377 | 34.6 (29.3-40.8) | - | - | - | 507 | 24.5 (20.2-29.8) | - | - | - | 318 | 35.1 (29.8-41.3) | - | - | - |
| Gender |  |  |  |  |  |  |  |  |  |  |  |  |  |  |  |
| Female | 277 | 35.7 (29.4-43.4) | reference | - | - | 370 | 24.2 (19.1-30.6) | reference | - | - | 223 | 36.1 (29.7-43.8) | reference |  |  |
| Male | 100 | 31.8 (23.3-43.6) | 0.89 (0.62-1.27) | 0.52 | 0.55 | 137 | 25.3 (17.8-36.0) | 1.05 (0.70-1.58) | 0.80 | 0.91 | 95 | 32.7 (23.9-44.5) | 0.91 (0.63-1.30) | 0.61 | 0.52 |
| Age at DM Diagnosis |  |  |  |  |  |  |  |  |  |  |  |  |  |  |  |
| <40 | 53 | 46.8 (31.4-69.9) | Reference | - |  | 66 | 44.9 (29.8-67.5) | Reference |  |  | 41 | 35.1 (22.1-55.7) | Reference |  |  |
| 40 – 50 | 124 | 34.0 (25.6-45.1) | 0.74 (0.45-1.20) | 0.22 | 0.39 | 172 | 27.6 (20.2-37.8) | 0.64 (0.38-1.06) | 0.08 | 0.35 | 120 | 40.4 (31.2-52.4) | 1.15 (0.67-1.95) | 0.61 | 0.32 |
| 50 – 60 | 132 | 33.7 (25.4-44.8) | 0.72 (0.44-1.18) | 0.19 | 0.44 | 178 | 18.9 (13.0-27.7) | 0.42 (0.24-0.73) | **0.002** | **0.05** | 109 | 35.8 (27.2-47.1) | 0.99 (0.58-1.69) | 0.98 | 0.49 |
| 60 – 70 | 52 | 31.4 (19.8-49.8) | 0.71 (0.40-1.27) | 0.25 | 0.62 | 73 | 15.7 (8.16-30.1) | 0.37 (0.18-0.78) | **0.009** | 0.137 | 41 | 24.4 (14.5-41.2) | 0.72 (0.36-1.45) | 0.36 | 0.89 |
| ≥70 | 16 | 18.8 (6.07-58.3) | 0.42 (0.12-1.45) | 0.17 | 0.33 | 18 | 12.5 (3.14-50.2) | 0.30 (0.07-1.30) | 0.107 | 0.965 | 7 | 18.8 (6.07-58.3) | 0.56 (0.18-1.81) | 0.34 | 0.69 |
| DM Duration (per 10 y) | 377 | - | 1.37 (1.10-1.72) | **0.005** | **0.03** | 507 | - | 2.00 (1.60-2.50) | **<0.01** | **<0.01** | 318 | - | 1.50 (1.23-1.84) | **<0.01** | **<0.01** |
| 0 – 5 y | 211 | 27.5 (21.2-35.7) | Reference | - |  | 272 | 14.7 (10.3-21.1) | Reference |  |  | 140 | 24.1 (18.2-31.8) | Reference |  |  |
| 5 – 10 y | 116 | 34.5 (25.6-46.6) | 1.24 (0.83-1.84) | 0.29 | 0.33 | 157 | 20.8 (14.2-30.7) | 1.37 (0.81-2.31) | 0.24 | 0.26 | 100 | 38.6 (29.1-51.2) | 1.58 (1.06-2.34) | **0.024** | **0.026** |
| >10 y | 50 | 52.7 (38.9-71.3) | 1.54 (1.04-2.31) | **0.03** | 0.09 | 78 | 55.2 (41.1-74.1) | 2.89 (1.82-4.57) | **<0.01** | **<0.01** | 78 | 57.7 (43.2-77.0) | 1.96 (1.31-2.95) | **0.001** | **0.002** |

P<sup>1</sup>: crude P<sup>2</sup>: multivariate regression model adjusted by age, gender, diabetes duration, systolic blood pressure, fasting blood glucose, gangrene, body mass index (BMI), diabetes medication, and residential area. Abbreviations: BMI, body mass index; CVD, cardiovascular diseases; DM, diabetes mellitus; DR, diabetic retinopathy; HS, high school; HR, hazard ratio; VTDR, vision threatening diabetic retinopathy

## Discussion

In this prospective cohort, we documented that every 1000 Indonesian adults with type 2 diabetes without DR, 34.6 persons would develop DR and among those already had DR, 35.1 persons would experience at least 1 step DR progression and 24.1 would develop VTDR every year. More importantly, 8.3 of these would be blind every year. We also identified that diabetes duration is the critical risk for the incidence and progression of DR and VTDR, whereas living in rural area is significant risk factor for blindness. These findings suggest that despite other factors, DR is progressing with increasing diabetes duration; thus, there is an urgent need to implement population-based screening program to early identify individuals who need treatment.

While there have been studies that published data about DR incidence, only very few of them reported the incidence rate of DR, particularly from developing countries [7]. This was the first evidence from Indonesian population showing DR incidence and progression. Incidence rate of DR in our population was lower compared to that reported by Tekele and colleagues in Ethiopia (48/100 person-years) [15]; but higher compared to a similar study in China, which had a DR incidence rate at 1.81/100 person-years [16]. Different population characteristics and disparities might contribute to these differences.

Despite the differences in incidence rate, we noted that longer diabetes duration was associated with increased risk of development and progression of DR, which is consistent with previous studies from Taiwan [17] and India [18]. Evidence has suggested that long-term hyperglycemia in diabetic individuals causes multi-faceted on-going biochemical and metabolic alterations leading to retinal microvascular injury [19,20]. The earlier the exposure affect some individuals – noted as younger diabetes diagnosis – or the longer the duration of diabetes, the greater the accumulation of these vicious changes, which are later followed by more prominent retinal microvascular damage [21–24]. We did not document significant associations of other risk factors such as blood glucose and blood pressure. We speculated that collective changes in various diabetes-related factors may be pertinent to long diabetes duration; thus, only diabetes duration was significantly associated with incidence and progression of DR in our study.

The strengths of this study include its prospective design, large sample size with high response rate, strict adherent to single examination protocol for baseline and follow-up, and side by side DR grading by a trained grader to ensure any retinal changes were documented. However, we also noted some limitations in this study. First, this follow-up was carried out at 5 years interval, therefore we lost exact time information of events that occurred in between examinations. Second, there was a chance of insufficient sample size because we only recruited sub-sample of the original cohort due to COVID-19 pandemic. Nevertheless, our associations, where existed, showed relatively narrow confidence interval, suggesting that we had sufficient study power to detect these associations.

In conclusion, our findings documented that every year, in every 1000 Indonesian adults with type 2 diabetes, 35 persons developed DR, 35 of those with DR got worse, 24 potentially lost their sight and 8 got blind. Individuals with diabetes duration of longer than 10 years had 1.5 times risk of developing DR and nearly triple the risk of vision loss within 5 years. These suggest that developing DR at some point over the course of diabetes might be inevitable. In the context of 21 millions of diabetes population in Indonesia in 2030 [3–5], there will be 500,000 individuals who potentially lose their vision and 168,000 new blindness every year if no action is taken. Therefore, nation-wide, promotive diabetes campaign to avoid early onset diabetes individuals and population-based eye screening program for persons with diabetes is mandatory to identify those needing timely treatment to prevent visual impairment and blindness.

## Data Availability

All relevant data are within the manuscript and its Supporting Information files

## Acknowledgments

The authors thank to the LHA Jogjakarta Province, and all CHCs for the technical assistance.

## Authors’ contribution

M.B.S. obtained funding, wrote, and substantially reviewed the manuscript, partial data analysis, contributed to study design. G.A.F. contributed to data collection and data management, partial data analysis and wrote the initial draft of the manuscript. Supanji contributed to grading verification, critically reviewed, and edited the manuscript. F.S.W contributed to grading verification, critically reviewed, and edited the manuscript. Y.D.L obtained funding, critically reviewed, and edited the manuscript. A.P. contributed to data collection, data checking, critically reviewed, and edited the manuscript. F.W contributed to research data interpretation, contributed to data collection, discussions, and critically reviewed and edited the manuscript.

## Supporting information

**S1 Table. Diabetic retinopathy incidence, VTDR incidence, and DR progression by other risk factors**

**S2 Table. Incidence rate of blindness**

